# Recent, Nearby Gun Violence and Recurrent Psychiatric Exacerbation in Children and Adolescents

**DOI:** 10.64898/2026.09.16.26362763

**Authors:** Carson S. Hartlage, Vivek Ashok, Erika Rasnick Manning, Meera Kotagal, Andrew F. Beck, Cole Brokamp

**Affiliations:** Department of Biostatistics, Health Informatics and Data Sciences, University of Cincinnati College of Medicine, Cincinnati, Ohio, United States; Division of Biomedical Informatics, Cincinnati Children’s Hospital Medical Center, Cincinnati, Ohio, United States; Department of Pediatrics, University of Cincinnati College of Medicine, Cincinnati, Ohio, United States; Division of General & Community Pediatrics, Cincinnati Children’s Hospital Medical Center, Cincinnati, Ohio, United States; Division of Biostatistics & Epidemiology, Cincinnati Children’s Hospital Medical Center, Cincinnati, Ohio, United States; Division of Pediatric General and Thoracic Surgery, Cincinnati Children’s Hospital Medical Center, Cincinnati, Ohio, United States; Department of Surgery, University of Cincinnati College of Medicine, Cincinnati, Ohio, United States; Division of Hospital Medicine, Cincinnati Children’s Hospital Medical Center, Cincinnati, Ohio, United States; Population Health, Cincinnati Children’s Hospital Medical Center, Cincinnati, Ohio, United States

## Abstract

**Importance:** Gun violence and youth mental illness are urgent public health concerns. Although prior studies have linked community violence with pediatric mental health outcomes, little is known about the connection between exposure to hyperlocal gun violence and the short-term risk of a psychiatric emergency at the individual level.

**Objective:** To estimate the impact of recent, nearby gun violence exposure on pediatric recurrent psychiatric exacerbation risk following an index psychiatric hospital admission or emergency department (ED) visit.

**Design:** Retrospective longitudinal cohort study using survival analyses with Cox proportional hazards models adjusted for season and census tract-level community deprivation and chronic crime.

**Participants:** Children and adolescents <18 years of age with a psychiatric hospital admission or ED visit at Cincinnati Children’s between January 1, 2016, and December 31, 2024, and a geocoded residential address within Cincinnati, Ohio.

**Exposure(s):** Recent, nearby gun violence, defined as at least one incident occurring within 400 meters of a patient’s geocoded residential addresses during the preceding 14 days.

**Main Outcome(s) and Measure(s):** Recurrent psychiatric exacerbation, defined as a subsequent psychiatric hospital admission or ED visit after an index exacerbation encounter.

**Results:** Among 4,730 children and adolescents (median age 13.7 years; [IQR, 10.6–15.7]; 54.1% female; 68.0% Black), participants contributed 2.1 million person-days of follow-up (median, 225 days [IQR, 38–654]). A total of 1,622 (34.3%) participants experienced a recurrent psychiatric exacerbation. Nearly half (41.5%) of participants were exposed to at least one nearby gun violence incident during follow-up. Exposure to gun violence within 400 meters of their home during the prior 14 days was associated with a higher hazard of recurrent psychiatric exacerbation in unadjusted (hazard ratio [HR], 2.26; 95% CI, 1.76–2.89) and adjusted (HR, 2.45; 95% CI, 1.91–3.14) models. Findings were robust across alternative spatial and temporal exposure definitions.

**Conclusions and Relevance:** Among youth with prior psychiatric emergency utilization, exposure to recent nearby gun violence was associated with a substantially increased risk of recurrent psychiatric exacerbation. Gun violence may represent an acute environmental trigger of psychiatric crises and a potentially actionable target for interventions to support vulnerable youth.

**KEY POINTS:** *Question:* Does recent exposure to nearby gun violence increase the risk of recurrent psychiatric exacerbation in youth with a history of acute psychiatric illness?

*Findings:* In this retrospective longitudinal cohort study of 4,730 children and adolescents, we found that gun violence within 400 meters of patients’ homes in the prior 14 days was associated with a significant increase in the hazard of recurrent psychiatric exacerbation.

*Meaning:* Gun violence may be an acute environmental precipitant for pediatric psychiatric crises, highlighting opportunities for intervention following incidents of gun violence and reinforcing the importance of efforts to prevent gun violence and its consequences.

## INTRODUCTION

Mental health-related emergency department (ED) visits and hospital admissions among US children and adolescents have increased substantially, and recurrent acute-care utilization is common.^1–3^ At the same time, gun violence is a major public health concern. It is the leading cause of death for children and adolescents aged 1-19,^4^ and its impact on child health and well-being extends beyond physical injury. Youth indirectly exposed to gun violence—including those who witness or hear an incident, live near an incident, or know someone who is injured or killed—may experience fear, loss, disruption of perceived safety, and traumatic stress.^5–11^ Gun violence exposure has also been linked to anxiety, depression, behavioral problems, and worse long-term mental health.^12–18^ One recent study found that neighborhood gun violence was associated with increased mental health-related ED utilization among children living nearby.^19^

Much of the existing literature has characterized gun violence over extended time periods, relied on area-level measures of exposure, or examined longer-term symptoms and health outcomes in broadly-defined pediatric populations. Although several studies have incorporated the distance between gun violence incidents and children’s homes or schools,^14–16,19,20^ and others have examined short-term, self-reported mental health symptoms after self-reported violence exposure,^21,22^ less is known about the immediate health impacts of discrete gun violence incidents occurring near a child’s residence. Specifically, it remains unclear whether recent, nearby gun violence may precipitate acute psychiatric crises among youth with pre-existing psychiatric conditions. Addressing this gap requires isolating short-term effects of gun violence exposure from chronic neighborhood conditions. Such evidence could clarify causal pathways and inform community- and healthcare-based interventions, including focused mental health supports following local incidents of gun violence.

To address this gap, we conducted a retrospective longitudinal cohort study evaluating the association between hyperlocal gun violence exposure and the hazard of emergency psychiatric encounters among children and adolescents in Cincinnati, Ohio, following an index psychiatric ED visit or emergency hospital admission. We hypothesized that recent, geographically proximate gun violence exposure would increase the risk of recurrent psychiatric exacerbation, independent of season, community material deprivation, and chronic neighborhood crime.

## METHODS

This study was reviewed and classified as exempt from ongoing review by the Cincinnati Children’s Hospital Medical Center (CCHMC) Institutional Review Board.

### Study setting and population

CCHMC is a quaternary pediatric academic medical center and one of the largest inpatient pediatric mental health providers in the US. CCHMC handles >95% of emergency pediatric psychiatric encounters within the city of Cincinnati, an urban area with a population of ∼314,500, including ∼65,100 children and adolescents.^23^

The study population included youth aged <18 years who had an emergency psychiatric encounter at CCHMC between January 1, 2016, and December 31, 2024, associated with a geocoded address in Cincinnati, Ohio. Encounters were eligible if they had one or more International Classification of Disease (ICD)-10 codes categorized in a psychiatric exacerbation category defined by the Pediatric Clinical Classification System (PECCS). We worked with a child and adolescent psychiatrist to refine PECCS categories a priori to retain codes with face validity for acute pediatric psychiatric emergencies based on current clinical practice and diagnostic formulation in the context of institutional coding and documentation practices. Exclusions were intended to improve specificity for acute pediatric emergency mental health encounters, recognizing that this may reduce sensitivity for broader behavioral health burden.^24^ Specific codes are provided in eTable 1.

### Constructing address histories

Patient-specific longitudinal residential address histories were constructed using encounters associated with a new residential address in the EHR. For the purpose of assessing exposure, moves were inferred as the temporal midpoint between encounters with different addresses, and residence was assumed to end 30 days after the last known new address. Patients were followed from the date of their first emergency psychiatric encounter during the study period. Follow-up was right censored at the earliest occurrence of the following: second psychiatric exacerbation during the study period, 18^th^ birthday, 30 days after last new address, or estimated move to an address outside Cincinnati.

### Outcome

The primary outcome was recurrent psychiatric exacerbation, defined as a second psychiatric-related ED visit or hospital admission after the first psychiatric exacerbation encounter within the study period.

### Exposure

Geocoded Gun Violence Archive (GVA) data for Hamilton County, Ohio, between January 1, 2016, and December 31, 2024, were downloaded on 2026-06-03 via The Trace’s Atlas of American Gun Violence.^25^ The GVA systematically collects and validates incidents of gun violence and gun crime, encompassing death, injury, threat, or use of a firearm, regardless of intent, outcome, or justification, using police, media, government, and other data sources.^26^ Suicides by gun (except murder-suicides), armed robberies without reported injuries, and incidents involving non-powder firearms (e.g., BB, airsoft) are excluded from this dataset.^26^ GVA records the best available location information, such as exact address, block (e.g., “1700 block of Vine St”), intersection (e.g., “Clark St and Cutter St”), or street (e.g., “Seymour Ave”), for each incident. The Trace maintains an interactive map of point- and day-level gun violence incidents using GVA data.^25^ Incidents with identical coordinates could have small, random spatial offsets applied, and incidents missing location information are excluded. Inspection of the raw GVA data for incidents tagged as Cincinnati during our study period (2016-2024) revealed 39 incidents without a precise location.

To define incidents relevant to our study, we restricted this dataset to incident points located within or up to 500m outside Cincinnati city boundaries. We included all other incidents regardless of outcome or type. We prespecified a primary exposure definition of recent, nearby gun violence exposure as one or more GVA incidents occurring within 400m of a patient’s current residence in the previous 14 days. This definition was chosen to reflect an acute exposure window based on existing literature and hypothesized short-term psychological stress responses.^19,21,22,27–30^ Incident coordinates were intersected with 400m buffered radii around patients’ geocoded residential addresses using the sf package in R and the NAD83 coordinate reference system. Figure 1A provides an example of the spatial buffer used to define exposure.

**Figure 1.**
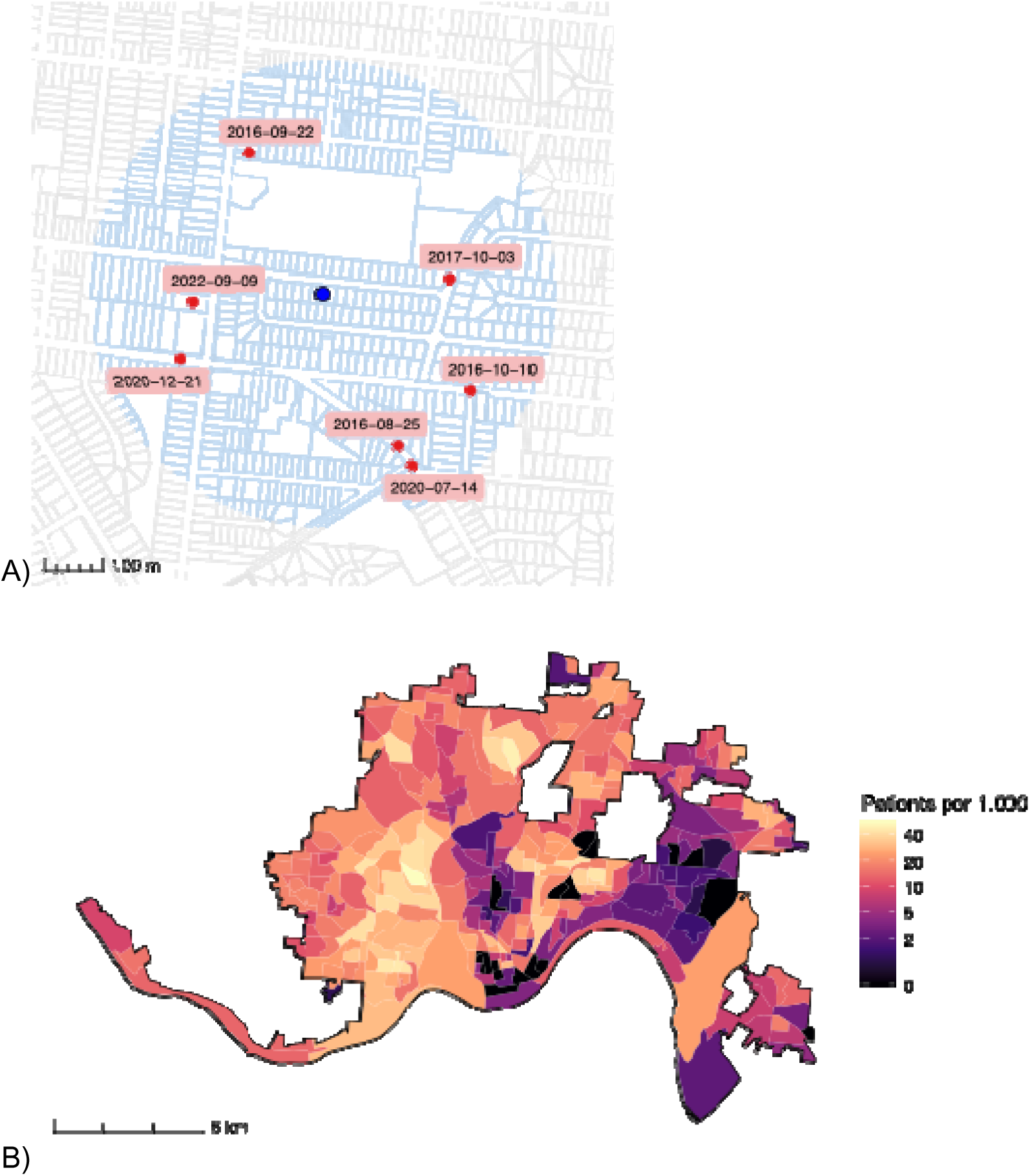
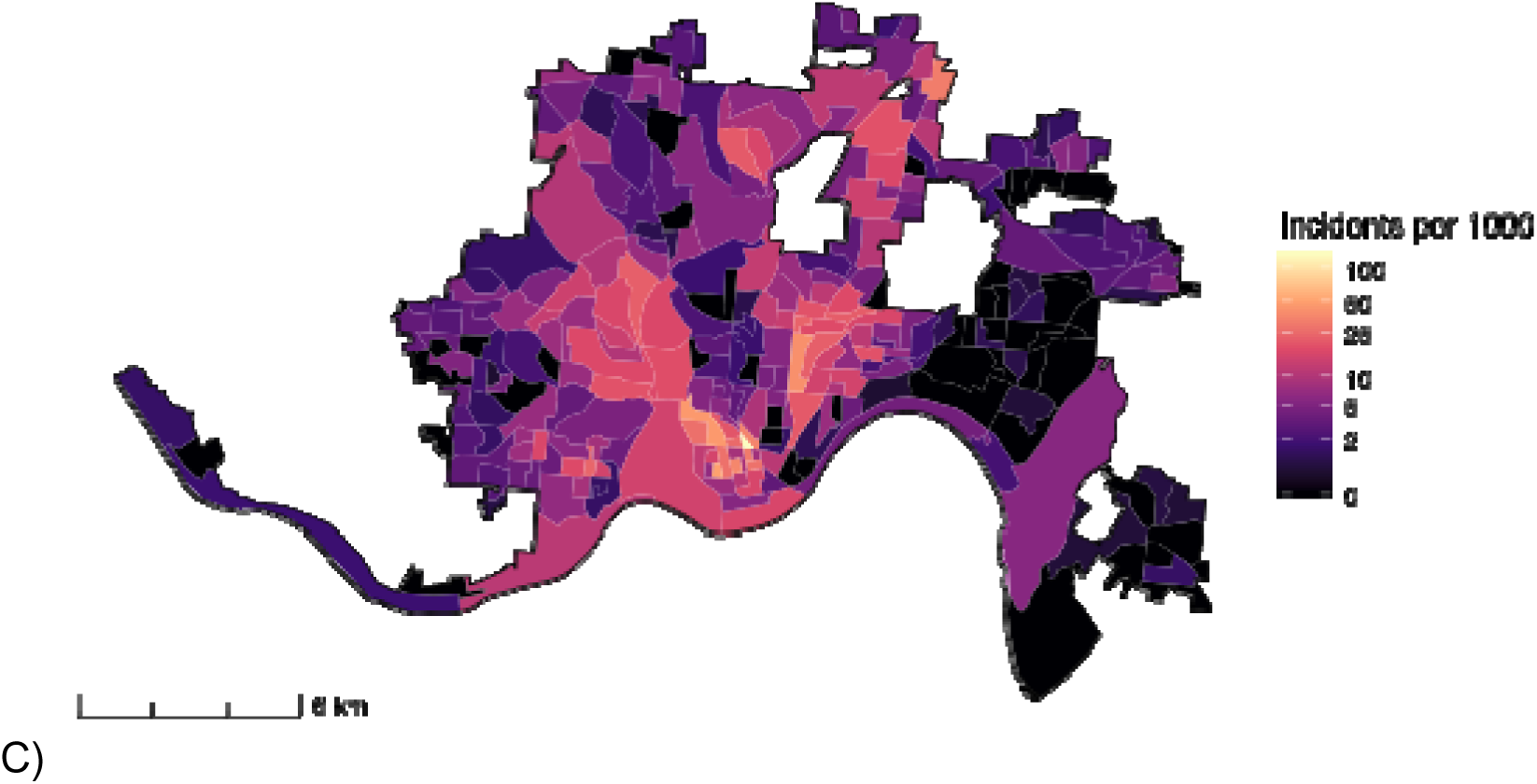
A) Example geocoded residential address in Cincinnati, Ohio, indicated by the blue dot, with a 400-meter radius buffer, indicated by the shaded blue circle. Gun violence incidents recorded by the Gun Violence Archive (2016-2024) and geocoded by The Trace are represented by red dots and labeled with dates. B) Cohort patients per 1,000 population by 2020 U.S. Census block group, based on each patient’s geocoded residential address at the time of inclusion. C) Gun violence incidents recorded by the Gun Violence Archive (2016-2024) and geocoded by The Trace in Cincinnati, Ohio, per 1,000 population by 2020 U.S. Census block group.

### Modeling approach

We used a Cox proportional hazards modeling approach with gun violence exposure treated as a time-varying exposure, using the counting-process (start–stop) formulation,^31^ to estimate associations between gun violence exposure and time to recurrent psychiatric exacerbation. To account for the correlation among repeated observations within individuals, robust standard errors were estimated using patient-level clustering. A Simon-Makuch plot was used to visualize the recurrent psychiatric exacerbation rate over time according to time-varying exposure status.^32^

The hypothesized impact of recent, nearby gun violence exposure on experiencing a recurrent psychiatric exacerbation was specified using a causal directed acyclic graph (DAG) developed based on literature review and expert input (Figure 2). The DAG identified three variables—season, community material deprivation, and chronic neighborhood crime—as sufficient covariates for blocking confounding when estimating the direct cause of recent, nearby gun violence exposure on recurrent psychiatric exacerbation. The data definitions used to represent these variables are provided in Table 1.

**Figure 2.**
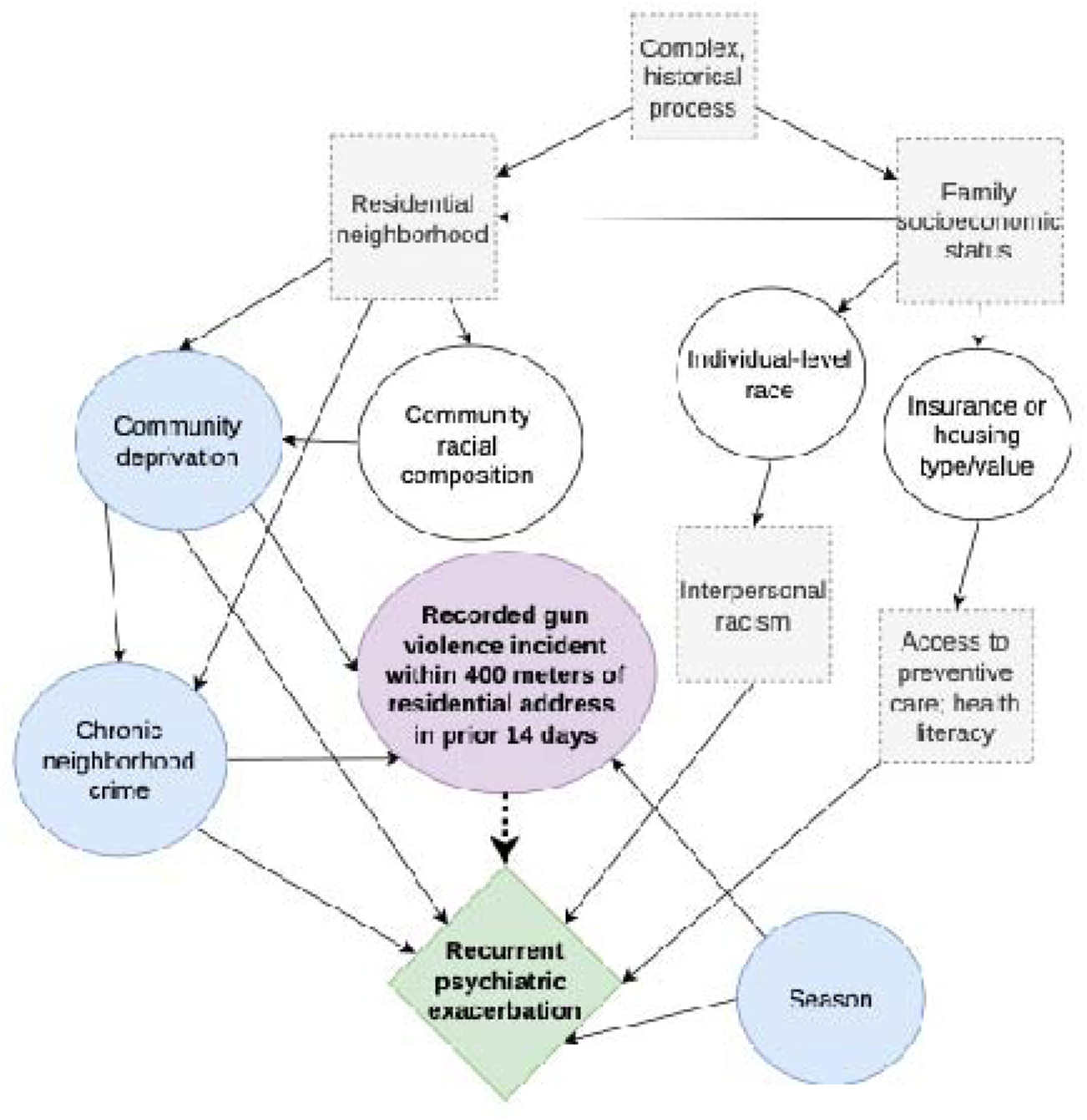
Directed acyclic graph illustrating the hypothesized relationships among exposure, outcome, and confounding variables. Green indicates the outcome, purple indicates the exposure, blue indicates measured confounding variables, white indicates non-confounding measured variables, and gray indicates unmeasured variables.

**Table 1.** Data definitions of exposure, outcome, and covariate constructs. Exposures and covariates were linked using patients’ longitudinal residential address histories.

| Construct | Variable type | Data used |
| --- | --- | --- |
| Recurrent psychiatric exacerbation | Outcome | Second psychiatric exacerbation encounter during the study period (2016-2024) |
| Recent, hyperlocal gun violence | Exposure | Day- and point-level incidents from Gun Violence Archive/The Trace, intersected with spatial buffers around patients' geocoded addresses |
| Community material deprivation | Covariate/confounder | Census tract-level Community Material Deprivation Index<sup>38</sup> constructed using 2023 American Community Survey variables |
| Chronic neighborhood crime | Covariate/confounder | Street-range violent and property crime incidents from the Cincinnati Police |
|  |  | Department<sup>39</sup> intersected with 2020 census tract geographies and summed over 2019-2023 |
| Season | Covariate/confounder (categorical) | Winter (12/1/XX-2/29/XX); Spring (3/1/XX-5/31/XX); Summer (6/1/XX-8/31/XX); Fall (9/1/XX-11/30/XX) |

Cox proportional hazards models were fit with these covariates to estimate adjusted hazards ratios (HRs). Because community deprivation and chronic neighborhood crime violated proportional hazards assumptions, they were converted to quartiles and included as strata. We estimated a model-based population attributable fraction using a counterfactual approach from the fitted Cox model.^33^ Specifically, we estimated the population-level risk of recurrent psychiatric exacerbation with the observed exposure histories and under a counterfactual scenario in which exposure was eliminated. The population attributable fraction was calculated as the difference between the average risks under these scenarios. We calculated the 95% confidence interval (CI) using 1,000 patient-level bootstrap resamples.

### Sensitivity analyses

We evaluated whether the observed association was robust to variations in exposure definitions, including smaller spatial radii (100m and 200m) and varying temporal windows (7, 21, 30, and 60 days). We fit unadjusted and adjusted Cox proportional hazards models for each distance-time exposure definition. We also evaluated secondary outcome definitions in which psychiatric exacerbation encounters occurring within 3 days and within 7 days of the index exacerbation were excluded as recurrent events, while patients continued to be followed thereafter. We did so because such encounters could have been part of the same exacerbation event.

## RESULTS

### Cohort characteristics

We followed 4,730 patients for 2,112,102 cumulative person-days. During follow-up, patients resided at 5,431 unique residential addresses across 272 census block groups (76.2% of total) in Cincinnati, Ohio (Figure 1B). The median follow-up time per patient was 225 days (interquartile range [IQR], 38–654), and 24.8% of patients had at least one address change during follow-up, with 1,769 observed address changes overall.

The median age at inclusion was 13.7 years (IQR, 10.6–15.7), and the cohort was mostly Black or African American (68.0%) and female (54.1%). Overall, 1,622 (34.3%) patients experienced a recurrent psychiatric exacerbation; compared to those who remained exacerbation-free, these patients were younger and more likely to be non-Hispanic but did not differ in sex, race, community deprivation, or chronic neighborhood crime (Table 2). Among patients who did not experience a recurrent exacerbation (n=3,108), 17.8% were right censored for age, and 82.2% were right censored for an estimated move outside of the city of Cincinnati or lack of updated address history.

**Table 2.** Cohort characteristics overall and by recurrent psychiatric exacerbation status. Continuous variables are displayed as median and interquartile range. P values were derived from Chi-squared tests for categorical variables, Wilcoxon rank-sum test for age, and t-tests for other continuous variables. Data definitions of community deprivation and chronic neighborhood crime are provided in Table 1.

| Characteristic | Overall (n=4,730) | No recurrent psychiatric exacerbation (n=3,108) | Recurrent psychiatric exacerbation (n=1,622) | p value |
| --- | --- | --- | --- | --- |
| Age at inclusion | 13.7 (10.6, 15.7) | 14.0 (10.4, 16.0) | 13.2 (10.7, 15.1) | **<0.001** |
| Sex: Female | 2558 (54.1%) | 1666 (53.6%) | 892 (55.0%) | 0.4 |
| Sex: Male | 2172 (45.9%) | 1442 (46.4%) | 730 (45.0%) |  |
| Ethnicity: Non-Hispanic | 4486 (94.8%) | 2931 (94.3%) | 1555 (95.9%) | **0.04** |
| Ethnicity: Hispanic | 201 (4.3%) | 149 (4.8%) | 52 (3.2%) |  |
| Ethnicity: Unknown | 43 (0.9%) | 28 (0.9%) | 15 (0.9%) |  |
| Race: White | 1119 (23.7%) | 718 (23.1%) | 401 (24.7%) | 0.1 |
| Race: Black or African American | 3215 (68.0%) | 2129 (68.5%) | 1086 (67.0%) |  |
| Race: Asian | 17 (0.4%) | 11 (0.4%) | 6 (0.4%) |  |
| Race: American Indian or Alaskan Native | 2 (0.0%) | 2 (0.1%) | 0 (0.0%) |  |
| Race: Native Hawaiian or Pacific Islander | 3 (0.1%) | 2 (0.1%) | 1 (0.1%) |  |
| Race: Multi-racial | 231 (4.9%) | 140 (4.5%) | 91 (5.6%) |  |
| Race: Unknown | 143 (3.0%) | 106 (3.4%) | 37 (2.3%) |  |
| Community deprivation | 0.41 (0.33, 0.56) | 0.41 (0.33, 0.56) | 0.41 (0.33, 0.56) | 0.8 |
| Chronic neighborhood crime | 1469 (847, 1924) | 1463 (838, 1886) | 1492 (873, 1946) | 0.5 |

### Exposure characteristics

Across the study area, 1,842 gun violence incidents occurred on 1,516 days during the study period (46.1% of all days) across 233 census block groups (63.3% of total; Figure 1C). Overall, 1,964 (41.5%) patients experienced an exposure to gun violence within 400m during follow-up, representing 1,472 incidents of gun violence (80.0% of all incidents) and 7,118 patient-level exposure instances. With a 14-day exposure window, 92,454 person-days (4.4% of all person-time) occurred during exposed periods. Among patients with any exposure, the median number of incidents experienced during follow-up was 2 (IQR, 1–4). Compared with patients without any exposure, those with at least one exposure lived in neighborhoods with higher material deprivation and chronic crime at the time of inclusion (eTable 2).

### Association between hyperlocal gun violence exposure and psychiatric re-exacerbation

Gun violence exposure within 400m of a patient’s residence and within 14 days was significantly associated with increased hazard of recurrent psychiatric exacerbation in the unadjusted model (HR, 2.26 [95% CI, 1.76–2.89]) and in the model adjusted for community deprivation, chronic neighborhood crime, and season (HR, 2.45 [95% CI, 1.91–3.14]). Figure 3 presents the Simon-Makuch curves for the probability of having a recurrent psychiatric exacerbation over time when exposed to gun violence.

**Figure 3.**
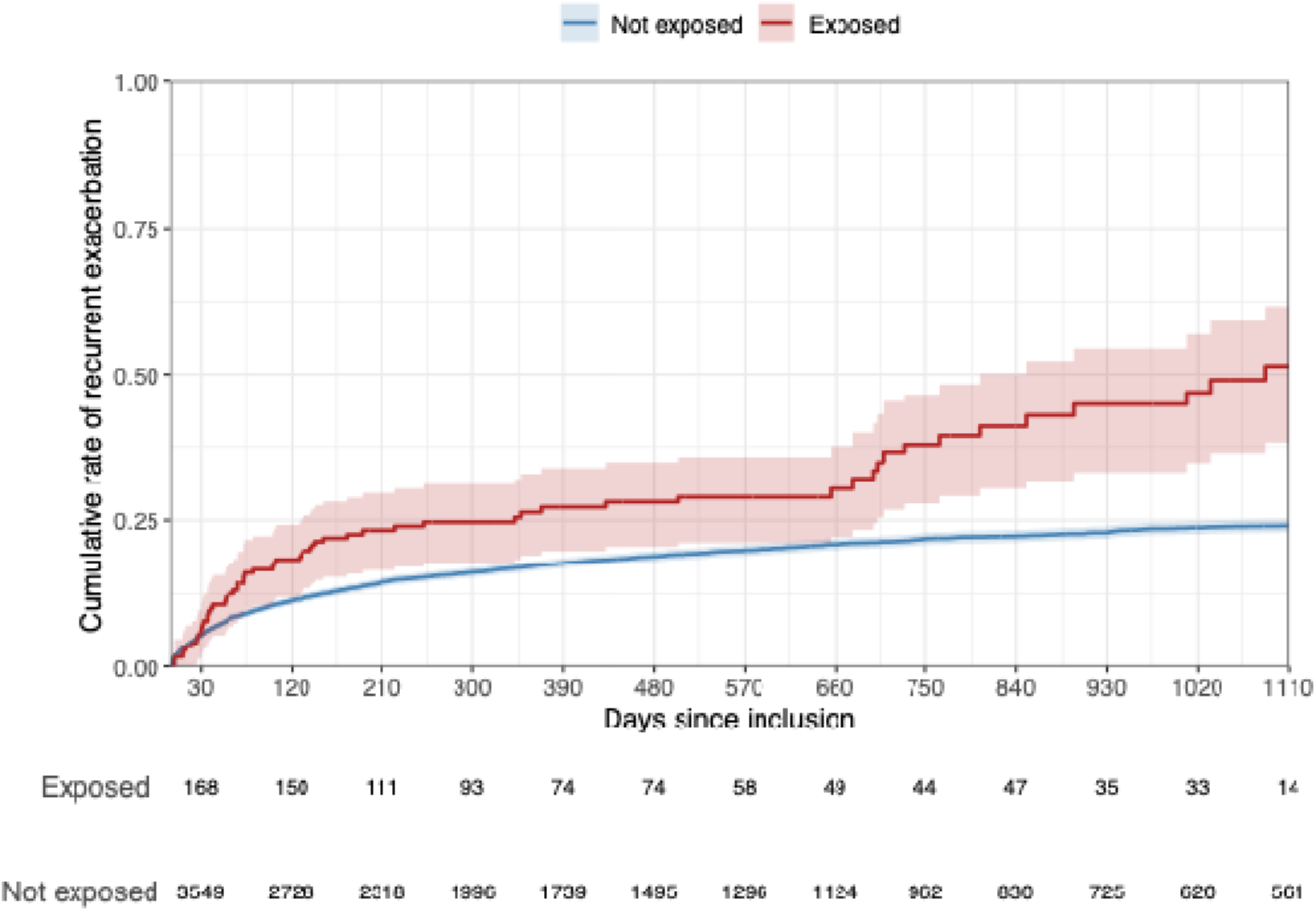
Simon-Makuch survival plot and risk table with exposure defined as any gun violence incident occurring within 400 meters of patient’s residential address in the prior 14 days. The plot’s x-axis was cropped to improve readability and includes the complete follow-up for 4,155 patients (87.8% of cohort). Shaded bands represent 95% confidence intervals.

The estimated population attributable fraction was 0.070 (95% CI, 0.035–0.099), suggesting that, under the assumptions of the causal model, eliminating gun violence in the study area during the study period could have reduced the population burden of recurrent psychiatric exacerbations by approximately 7.0%, corresponding to an estimated 114 fewer exacerbations.

Results were generally robust across alternative spatial and temporal exposure definitions (eTable 3). Both 100m and 200m exposure definitions showed significant associations between gun violence exposure and psychiatric exacerbation. Effect estimates for the 400m definition were larger than other spatial radii and attenuated with increasing time beyond 14 days.

Additionally, results for the primary exposure definition were consistent when outcomes within the first 3 days of follow-up were suppressed (98 exacerbation encounters excluded; unadjusted HR, 2.29 [95% CI, 1.79–2.94]; adjusted HR, 2.48 [95% CI, 1.94–3.18]) and when outcomes within the first 7 days of follow-up were suppressed (168 exacerbation encounters excluded; unadjusted HR, 2.31 [95% CI, 1.79–2.96]; adjusted HR, 2.50 [95% CI, 1.95–3.20]).

## DISCUSSION

In a longitudinal cohort of youth with a history of psychiatric emergency care utilization, we found a significant increase in the hazard of a recurrent psychiatric exacerbation in the 14 days following a shooting within 400m of a child’s residence. This association persisted with adjustment for season, community deprivation, and chronic neighborhood crime and was robust across alternative exposure and outcome definitions. These findings suggest that the pediatric psychiatric effects of indirect exposure to gun violence may emerge within days to weeks and be severe enough to result in a need for emergency care, independent of long-term adverse neighborhood conditions.

Our findings are consistent with prior studies linking gun violence and pediatric mental health.^7–9^ For example, in nationwide samples, Buggs et al. and Leibbrand et al. found associations between firearm homicide near (within 1300m and 1 mile, respectively) youths’ homes or schools, and worse mental and behavioral health outcomes within one year.^14,15^ Similarly, Gard et al. reported that deadly gun violence occurring within 500m of youths’ homes in the prior year was associated with increased behavioral problems and demonstrated both spatial and cumulative dose-response relationships.^16^ In a Philadelphia-based study, Vasan et al. found increased mental health-related ED utilization among children living within ¼ mile of a shooting in the subsequent 60 days.^19^ They also observed larger effect sizes at a ⅛ mile radius, and the strongest temporal association was found to be within 2 weeks after a shooting.^19^ Relatedly, two studies of self-reported violence exposure, rather than gun violence specifically, found increases in youths’ mental health symptoms on the same or following day.^21,22^

We build on this literature by identifying an acute and potentially actionable period of elevated psychiatric risk following a local gun violence event. Specifically, our study extends prior work by examining patient-level longitudinal risk among children with pre-existing psychiatric vulnerability rather than changes in population-level utilization rates or long-term self-reported symptoms. We also show that the effects of nearby gun violence may not be solely attributable to the cumulative burden of living in a materially deprived or high-crime neighborhood; it may act as acute environmental precipitant that triggers psychiatric exacerbations within weeks rather than months or years.

Several mechanisms may explain this relationship. Even when children are not directly victimized by gun violence, an incident may heighten vigilance and anxiety, disrupt feelings of safety, and trigger traumatic stress responses.^21,22^ Hyperlocal gun violence may also increase caregiver stress, strain social networks, and contribute to changes in school attendance and outdoor activity.^34,35^ For youth with pre-existing psychiatric conditions, these stressors could overwhelm coping capacities and precipitate psychiatric symptom escalation. Our finding of increased risk within the two weeks following a nearby gun violence incident is consistent with these hypothesized short-term psychosocial and behavioral disruptions, suggesting that the immediate aftermath may represent a critical period of vulnerability.

Our study has several strengths. First, gun violence was modeled as a time-varying, spatiotemporally precise exposure, improving upon approaches relying solely on cross-sectional or spatially- or temporally-aggregated measures. Further, we used an objective measure of reported gun violence that could not be impacted by observational or recall biases usually present in self-reported exposures. Adjustment for community deprivation and chronic neighborhood crime enabled us to isolate the contribution of acute gun violence events beyond persistent neighborhood disadvantage. Additionally, because CCHMC is the dominant provider of emergency psychiatric care in our region, records provided a large cohort and population-wide capture of our outcome.

Several limitations should be considered when interpreting these findings. First, the study was conducted in a single city, limiting generalizability to non-urban settings or regions with different demographic composition or patterns of gun violence and healthcare utilization. Second, our cohort was restricted to youth with a prior psychiatric emergency encounter. Although this population was intentionally selected because it was hypothesized to be particularly susceptible to acute environmental stressors, the findings may not generalize to broader populations without prior established psychiatric morbidity. Further, exposure misclassification is possible. Although GVA is a valuable open resource of national data, it may incompletely capture gun violence incidents or contain geographic inaccuracies. Also, residential proximity may or may not reflect an individual’s awareness of an incident, whether they witnessed or heard it, or whether they personally knew someone involved. In addition, longitudinal residential histories were inferred from EHR addresses, whose accuracy is impacted by engagement with the healthcare system. Further, youth spend substantial time in settings outside recorded residential addresses, including schools, childcare facilities, and homes of other caregivers, potentially leading to under- or overestimation of true exposure. Finally, as with all observational studies, residual confounding from unmeasured individual-, family-, or neighborhood-level factors remains possible.

Our findings have important implications for clinical practice, public health, and policy. Unlike many social and environmental drivers of mental health that are diffuse and difficult to react to in a timely manner, nearby shootings represent easily identifiable events that may signal elevated risk of psychiatric crisis among vulnerable youth. The period following a shooting may therefore constitute a critical intervention window during which targeted support could reduce the need for emergency psychiatric care. Healthcare systems already possess the geocoded addresses and psychiatric care utilization information necessary to identify children at risk. Public gun incident databases like GVA could be integrated into population health infrastructure on a daily or weekly basis to trigger targeted outreach following local shootings. Such outreach might include care coordination, telehealth behavioral health check-ins, school-based supports, proactive crisis hotline engagement, or services directed toward caregivers. For example, qualitative work with African American youth living in low-income urban neighborhoods has identified unmet needs for access to mental health services following experiences of community violence.^36^ Hospital-based violence intervention programs, which provide trauma-informed services to youth after violent injury, represent an analogous approach to focused support and prevention following violence exposure.^37^

Future studies should evaluate whether these findings are reproducible across other geographic settings, pediatric populations, and healthcare systems. Additional work is also needed to better characterize dose- and lag-response relationships, including the effects of exposure proximity, frequency, and cumulative exposure over time. Further research should investigate potential mechanisms underlying these associations and whether certain psychiatric outcomes, such as suicidality, anxiety disorders, or trauma-related disorders, are particularly sensitive to nearby shootings. Finally, intervention studies and simulation modeling could help assess the feasibility, effectiveness, and cost-effectiveness of screening, outreach, surveillance strategies, and policies designed to reduce violence-attributable psychiatric morbidity among youth.

## CONCLUSION

Among children and adolescents with pre-existing psychiatric vulnerability, exposure to a nearby gun violence incident was associated with an increased hazard of psychiatric emergency care utilization within weeks. Recognizing gun violence as a time-sensitive mental health risk factor may inform both violence-prevention efforts and targeted interventions designed to support youth following local shooting incidents.

## Data Availability

Gun violence incident data, community material deprivation index, street-range crime incident data, and analytic code are publicly available online at the links provided. The healthcare data used in this article cannot be shared because they contain protected health information under HIPAA and were used under an IRB-approved research study with exempt use of protected health information and no sharing beyond the study investigators.

https://projects.thetrace.org/gun-violence-map/?state=Ohio&locationType=county&location=Hamilton+County

https://github.com/geomarker-io/dep_index

https://data.cincinnati-oh.gov/safety/PDI-Police-Data-Initiative-Crime-Incidents/k59e-2pvf/about_data

https://github.com/carson-hartlage/gv-rpe-manuscript

## ACKNOWLEDGEMENTS

We are grateful to the teams at the Gun Violence Archive, The Trace, and the City of Cincinnati Office of Performance & Data Analytics for their commitment to collecting, maintaining, and sharing data. This work was supported by the National Institute of Environmental Health Sciences (R03ES037996) and the Agency for Healthcare Research and Quality (R01HS027996). The authors have no potential conflicts of interest to disclose.

## Author contributions

Carson Hartlage and Cole Brokamp had access to all data used in the study and take responsibility for the integrity of the data and the accuracy of the data analysis.

Concept and design: Hartlage, Beck, Brokamp.

Acquisition, analysis, or interpretation of data: All authors.

Drafting of the manuscript: Hartlage, Brokamp.

Critical revision of the manuscript for important intellectual content: All authors.

Statistical analysis: Hartlage, Brokamp.

Administrative, technical, or material support: Manning, Beck, Brokamp.

Supervision: Beck, Brokamp.

