## Supplement for "Recent, Nearby Gun Violence and Recurrent Psychiatric Exacerbation in Children and Adolescents"

**ONLINE-ONLY SUPPLEMENT**

**Psychiatric exacerbation definition**

We excluded selected F-codes that represented mental disorders due to known physiological or medical conditions (e.g., F06.4, F06.30, F06.0), chronic or outpatient-oriented conditions unlikely to drive an ED visit (e.g., F30.1, F21), residual or nonspecific diagnostic categories (e.g. F40.9), low-prevalence or adult-oriented impulse-control conditions (e.g., F91.0, F63.0), context-specific behavioral diagnoses, (e.g., F42.2, F42.3, F42.4) and remission or mild-state bipolar disorder codes (e.g., F31.11). Examples of excluded codes for impulse control disorders: "pathological gambling", "kleptomania", "trichotillomania"; for anxiety disorders: "hoarding disorder", "excoriation disorder", "nonpsychotic mental disorder"). Included billing codes are summarized in eTable 1.

**eTable1.** Select billing codes used to define pediatric psychiatric exacerbations grouped by the corresponding PECCS category. PECCS: Pediatric Clinical Classification System; ICD: International Classification of Diseases.

| **PECCS category** | **ICD-10-CM codes** |
| --- | --- |
| Adjustment disorders | F43.20, F43.21, F43.22, F43.23, F43.24, F43.25, F43.29, F43.8, F43.9 |
| Anxiety disorders | F41.0, F41.1, F41.3, F41.8, F41.9, F43.0 |
| Disorders usually diagnosed in infancy childhood or adolescence | F93.0 |
| Attention-deficit hyperactivity disorder | F90.0, F90.1, F90.2, F90.8, F90.9 |
| Conduct disorder | F91.1, F91.2, F91.8, F91.9 |
| Oppositional defiant disorder | F91.3 |
| Disruptive behavior disorder | R46.89 |
| Impulse control disorders NEC | F63.1, F63.89, F63.9, R45.850 |
| Intermittent explosive disorder | F63.81 |
| Mood disorders | F34.0, F34.1, F34.8, F34.81, F34.89, F34.9, F39 |
| Mood disorders (bipolar disorder) | F31.0, F31.10, F31.12, F31.13, F31.2, F31.30, F31.31, F31.32, F31.4, F31.5, F31.60, F31.61, F31.62, F31.63, F31.64, F31.70, F31.73, F31.75, F31.77, F31.81, F31.89, F31.9 |
| Mood disorders (major depressive disorder) | F32.0, F32.1, F32.2, F32.3, F32.5, F32.8, F32.89, F32.9, F33.0, F33.1, F33.2, F33.3, F33.40, F33.8, F33.9 |
| Mood disorders (manic episodes) | F30.2, F30.8, F30.9 |
| Personality disorders | F60.3, F60.89, F60.9 |
| Phobias | F40.00, F40.01, F40.10, F40.11, F40.298 |
| Posttraumatic stress disorder | F43.10, F43.11, F43.12 |
| Schizophrenia and other psychotic disorders | F20.0, F20.1, F20.2, F20.3, F20.5, F20.81, F20.89, F20.9, F22, F23, F24, F25.0, F25.1, F25.8, F25.9, F29 |
| Suicide and intentional self-inflicted injury | R45.851, T14.91, T36.1X2A, T37.5X2A, T38.1X2A, T38.3X2A, T39.012A, T39.1X2A, T39.1X2D, T39.312A, T39.392A, T39.8X2A, T39.92XA, T40.2X2A, T40.4X2A, T40.7X2A, T40.8X2A, T42.4X2A, T42.6X2A, T42.8X2A, T43.012A, T43.212A, T43.222A, T43.292A, T43.3X2A, T43.592A, T43.612A, T43.622A, T43.632A, T44.3X2A, T44.7X2A, T44.992A, T45.0X2A, T45.2X2A, T45.4X2A, T46.1X2A, T46.4X2A, T46.5X2A, T47.0X2A, T47.92XA, T48.1X2A, T48.3X2A, T48.4X2A, T49.0X2A, T50.902A, T50.992A, T51.0X2A, T54.1X2A, T54.92XA, T65.892A, T71.162A |

**eTable 2.** Cohort characteristics overall and by any gun violence exposure within 400 meters during follow-up status. Continuous variables are displayed as median and interquartile range. P values were derived from Chi-squared tests for categorical variables, Wilcoxon rank-sum test for age, and t-tests for other continuous variables. Data definitions of community deprivation and chronic neighborhood crime are provided in Table 1.

| **Characteristic** | **Overall (n=4,730)** | **No gun violence exposure during follow-up (n=2,766)** | **Exposure to at least one gun violence incident during follow-up (n=1,964)** | **p value** |
| --- | --- | --- | --- | --- |
| Age at inclusion | 13.7 (10.6, 15.7) | 14.2 (11.4, 16.0) | 12.9 (9.8, 15.1) | **<0.001** |
| Sex: Female | 2558 (54.1%) | 1507 (54.5%) | 1051 (53.5%) | 0.5 |
| Sex: Male | 2172 (45.9%) | 1259 (45.5%) | 913 (46.5%) |  |
| Ethnicity: Non-Hispanic | 4486 (94.8%) | 2610 (94.4%) | 1876 (95.5%) | 0.06 |
| Ethnicity: Hispanic | 201 (4.3%) | 124 (4.5%) | 77 (3.9%) |  |
| Ethnicity: Unknown | 43 (0.9%) | 32 (1.1%) | 11 (0.6%) |  |
| Race: White | 1119 (23.7%) | 796 (28.8%) | 323 (16.4%) | **<0.001** |
| Race: Black or African American | 3215 (68.0%) | 1714 (62.0%) | 1501 (76.4%) |  |
| Race: Asian | 17 (0.4%) | 15 (0.5%) | 2 (0.1%) |  |
| Race: American Indian or Alaskan Native | 2 (0.0%) | 0 (0.0%) | 2 (0.1%) |  |
| Race: Native Hawaiian or Pacific Islander | 3 (0.1%) | 2 (0.1%) | 1 (0.1%) |  |
| Race: Multi-racial | 231 (4.9%) | 145 (5.2%) | 86 (4.4%) |  |
| Race: Unknown | 143 (3.0%) | 94 (3.4%) | 49 (2.5%) |  |
| Community deprivation | 0.41 (0.33, 0.56) | 0.40 (0.33, 0.56) | 0.47 (0.36, 0.59) | **<0.001** |
| Chronic neighborhood crime | 1469 (847, 1924) | 1353 (790, 1810) | 1604 (985, 2028) | **<0.001** |

**eTable 3.** Results for Cox proportional hazards models fit using alternative exposure definitions. 13.1% of patients had any exposure within 100m during follow-up, and 27.5% had any exposure within 200m. HR: hazard ratio; CI: confidence interval

| **Distance** | **Time** | **Unadjusted HR (95% CI)** | **Adjusted HR (95% CI)** |
| --- | --- | --- | --- |
| 100m | 7d | 1.28 (0.53 - 3.07) | 1.39 (0.58 - 3.34) |
| 100m | 14d | 1.65 (0.93 - 2.92) | 1.74 (0.98 - 3.08) |
| 100m | 21d | 1.73 (1.09 - 2.76) | 1.78 (1.11 - 2.85) |
| 100m | 30d | 1.34 (0.85 - 2.11) | 1.37 (0.86 - 2.17) |
| 100m | 60d | 1.13 (0.78 - 1.65) | 1.17 (0.80 - 1.71) |
| 200m | 7d | 1.30 (0.74 - 2.31) | 1.46 (0.83 - 2.57) |
| 200m | 14d | 1.43 (0.97 - 2.13) | 1.57 (1.06 - 2.34) |
| 200m | 21d | 1.39 (0.99 - 1.94) | 1.51 (1.08 - 2.11) |
| 200m | 30d | 1.28 (0.95 - 1.72) | 1.38 (1.02 - 1.88) |
| 200m | 60d | 1.18 (0.93 - 1.50) | 1.27 (0.99 - 1.62) |
| 400m | 7d | 2.08 (1.46 - 2.96) | 2.27 (1.60 - 3.22) |
| 400m | 14d | 2.26 (1.76 - 2.89) | 2.45 (1.91 - 3.14) |
| 400m | 21d | 2.21 (1.78 - 2.73) | 2.40 (1.94 - 2.97) |
| 400m | 30d | 2.03 (1.68 - 2.46) | 2.19 (1.81 - 2.66) |
| 400m | 60d | 1.88 (1.61 - 2.19) | 2.03 (1.74 - 2.37) |
